# Detection of a BA.1/BA.2 recombinant in travelers arriving in Hong Kong, February 2022

**DOI:** 10.1101/2022.03.28.22273020

**Authors:** Haogao Gu, Daisy YM Ng, Gigi YZ Liu, Samuel SM Cheng, Pavithra Krishnan, Lydia DJ Chang, Sammi SY Cheuk, Mani MY Hui, Tommy TY Lam, Malik Peiris, Leo Poon

## Abstract

We studied SARS-CoV-2 genomes from travelers arriving in Hong Kong from November-2021 to February-2022. Apart from detecting Omicron (BA.1, BA1.1. and BA.2) and Delta variants, we detected a BA.1/BA.2 recombinant in two epidemiologically linked cases. This recombinant has a breakpoint near the 5’ end of Spike gene (nucleotide position 20055-21618).

## Main text

The SARS-CoV-2 Omicron variant (Pango lineage: B.1.1.529) emerged in November-2021. Three subvariants of Omicron (BA.1/BA.1.1/BA.2) were detected in varying proportion in different continents within weeks, with BA.1 being initially the dominant (1). As of March-2022, these 3 subvariants accounted for >95% of the sequences submitted to GISAID. We previously demonstrated the feasibility of using incoming travelers for SARS-CoV-2 genomic surveillance (2). Here, we report the detection of a BA.1/BA.2 recombinant in travelers through our surveillance.

Using our previously described next-generation sequencing method (2), we analyzed 198 out of 793 (25%, C_t_ values:<30) SARS-CoV-2 RT-PCR positive samples collected from travelers arriving in Hong Kong from 15-November-2021 to 4-February-2022 (Appendix Table 1 and Appendix Methods). We successfully deduced near-full genome sequences from 180 samples. The deduced genomes predominantly belonged to Delta (N=58) and Omicron (BA.1=66, BA.1.1=28 and BA.2=26) variants (Appendix Figures 1 and 2). The time distribution of these variants agrees with the global surveillance data submitted to GISAID (https://www.gisaid.org/phylodynamics/global/nextstrain/), confirming that travel hubs are useful sentinel sites to monitor SARS-CoV-2 circulation (2). Interestingly, our BA.2 cases predominantly were imported from Philippines and Nepal, indicating that this subvariant might have established itself in these countries early.

In our phylogenetic analysis, two additional nearly identical sequences formed a distinct branch in the Omicron clade (Appendix Figure 2; Arrow). These genomes were detected from two epidemiologically linked cases (Patients 1 and 2). These patients travelled together to Hong Kong in February-2022 from Europe. They were RT-PCR tested positive for SARS-CoV-2 at airport upon arrival (C_t_ values: 27 and 22). Patient 1 reported to have sore throat and cough in late January-2022, whereas Patient 2 was asymptomatic. Both patients had received two doses of COVID-19 vaccine (Pfizer-BioNTech), with the second dose given in Nov-2021 and June-2021 for Patient 1 and Patient 2, respectively.

The distinct topology of viral sequences from Patients 1 and 2 prompted us to hypothesize that they were infected by a recombinant virus. To test this hypothesis, we used previously reported BA.1- and BA.2-defining single-nucleotide polymorphisms (SNPs) to analyze these genomes (Appendix Methods). As shown in Figure A, the 5’ end sequences (nucleotide region: 1-20055) of these 2 cases only contain BA.1-specific SNPs. By contrast, their corresponding 3’ end sequences only contain BA.2-specific SNPs. We further conducted a recombination analysis and confirmed that there is only one breakpoint located between nucleotide positions 20055-21618 in this recombinant (Appendix Figure 3). The nucleotide sequences 5’ and 3’ to this breakpoint are phylogenetically similar to authentic BA.1 and BA.2 sequences, respectively (Figure B). The breakpoint identified in this recombinant virus locates near the 5’ ORF of spike gene. Recombinant viruses (e.g. B.1.1.7/B1.177 and Delta/BA1) with a breakpoint in this region were also reported by others (3) (https://www.medrxiv.org/content/10.1101/2022.03.03.22271812v2 and https://github.com/cov-lineages/pango-designation/issues/441), suggesting that this region might be a hotspot for recombination.

**Figure 1.**
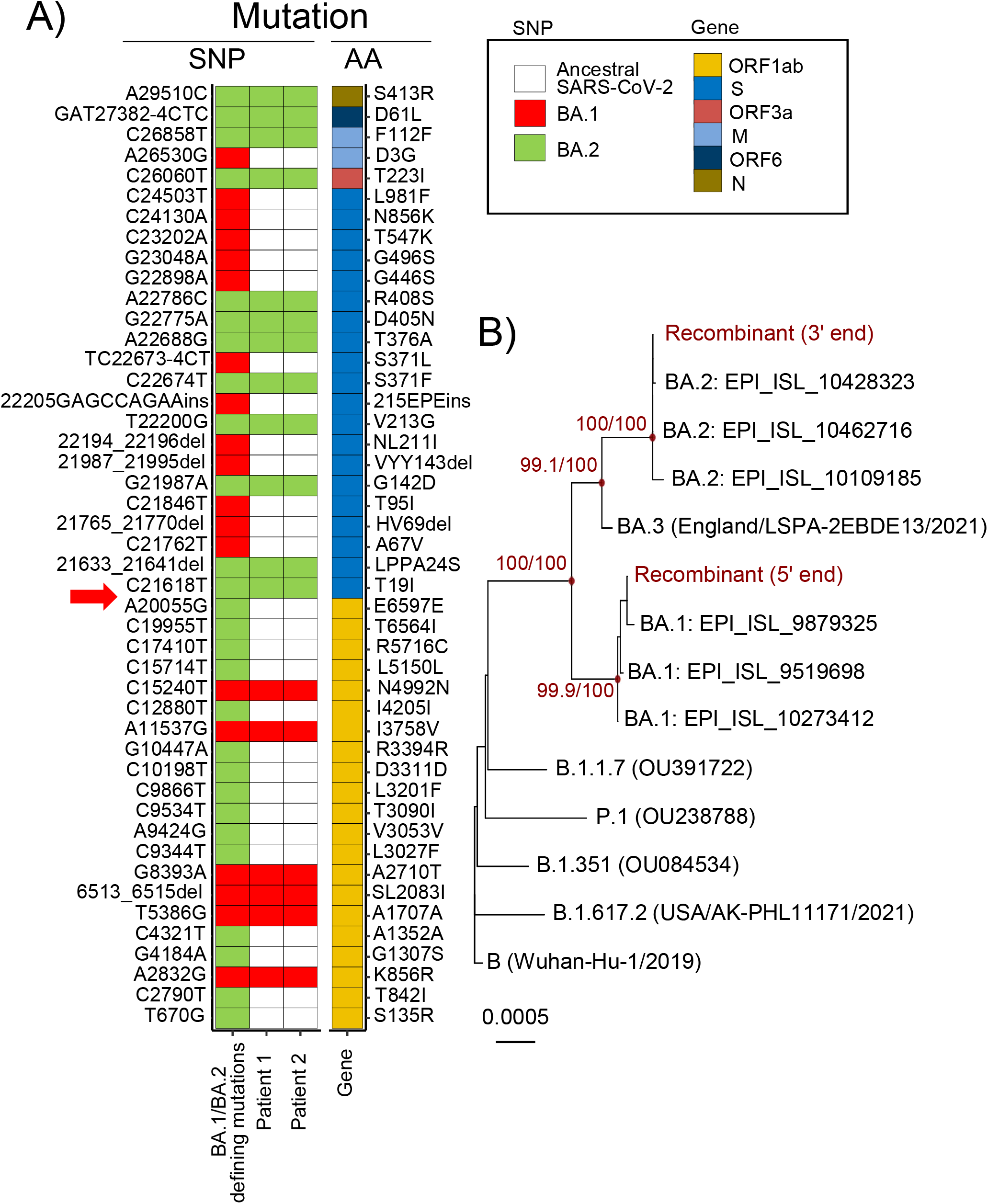
Detection of a BA.1/BA.2 recombinant. A) Mapping of BA.1- and BA.2-specific SNPs with respect to the reference sequence (Genbank: MN908947.3). BA.1- and BA.2-specific SNPs found in Patients 1 and 2 samples are shown in red and blue boxes, respectively. The corresponding amino acid changes of these SNPs are also indicated. The putative breaking point is indicated by an arrow. B) Phylogeny of viral RNA sequences 5’ and 3’ to the putative breaking point. The Maximum likelihood tree was generated using IQ-TREE employing the TIM+F+I nucleotide substitution model with Wuhan-Hu-1 (GenBank: MN908947.3) as the outgroup. References sequences, together with their accession numbers in GISAID or Genbank, are shown as indicated. The red node points show strongly supported branches by SH-aLRT and ultrafast bootstrap values. Scale bar indicates estimated genetic distance.

We further examined our sequencing data to exclude the possibility of co-infection or contamination (4). We noted that the minor allele frequencies (MAF) at these BA.1- and BA.2-defining SNP positions were extremely low (Figure A, Mean=0.5%, Median=0.06%), indicating that these samples only contained a single virus population. Besides, we used Patient 2 sample to clone a RT-PCR product (∼2.2k bp) spanning the recombination breakpoint. Both BA.1-specific (C19955T/A20055G) and BA.2-specific (21633-21641del/C21762T) SNPs can be detected in the same plasmid clone, confirming that Patients 1 and 2 were indeed infected by a BA.1/BA.2 recombinant virus.

We did not find similar BA.1/BA.2 recombinant sequences in GISIAD or Genbank (accessed on 7-March-2022), suggesting this is a novel recombinant. The BA.1 region of this recombinant virus is genetically closed to 3 BA.1 sequences detected in Europe and USA (Fig. B), whereas its BA.2 region is identical to a vast number of BA.2 sequences from multiple continents (N=19,555). With such a high global co-circulation of BA.1 and BA.2 subvariants, it would be difficult to pinpoint the geographical location where this recombination event occurred.

The emergence of Omicron subvariants that allow vaccine breakthrough and widespread re-infection is of public health concern. Previous studies reported the detection of SARS-CoV-2 interlineage recombinants at the time when different SARS-CoV-2 lineages were co-circulating (3,5) (https://www.medrxiv.org/content/10.1101/2022.03.03.22271812v2 and https://github.com/cov-lineages/pango-designation/issues/441). The high transmissibility of Omicron (6,7) have led to a wide co-circulation of BA.1 and BA.2 subvariants in many regions. This might provide ample opportunities for them to generate novel recombinants, either amongst themselves or with other variants, via co-infection events. Although the current global surveillance data suggest that our recombinant might only be a sporadic case, the potential impacts of this kind of novel recombinants should not be underestimated. One should note that homologous recombination is very common in animal and other human coronaviruses (8) and that some of these recombination events could generate recombinants with enhanced virulence (9,10). This urges for the need of a long-term genomic surveillance of SARS-CoV-2 at global level.

## Biography

Haogao Gu is a postdoctoral fellow at The University of Hong Kong, Hong Kong. His interests focus on bioinformatics and virus evolution.

## Data Availability

All deduced sequences can be available from GISAID.
The GISAID accession number of our sequences are available online at:
https://github.com/Leo-Poon-Lab/BA1_BA2_recombinant_HK/blob/main/GISAID_accessions.txt

## Ethics

The work is approved by IRB at the University of Hong Kong (IRB number: UW 20-168).

## Acknowledgements

We gratefully acknowledge the staff from the originating laboratories responsible for obtaining the specimens and from the submitting laboratories where the genome data were generated and shared via GISAID (Appendix Table 2). We acknowledge the technical support provided by colleagues from the Centre for PanorOmic Sciences of the University of Hong Kong. This work is supported by Research Grants Council of Hong Kong theme-based research schemes (T11-705/21-N) and the Health and Medical Research Fund (COVID190205).

We also acknowledge the Centre for Health Protection of the Department of Health for providing epidemiological data for the study.

