## Appendix methods, figures and tables for "Detection of a BA.1/BA.2 recombinant in travelers arriving in Hong Kong, February 2022"

### *Sequencing*

Clinical samples from SARS-CoV-2 RT-PCR tested positive case-patients (Ct value <30) were subjected to next generation sequencing. RNA samples were sent to a WHO reference laboratory at the University of Hong Kong for full genome analyses (IRB number: UW 20-168). We deduced near full-length genomes from the samples using a Illumina sequencing protocol previously described by us (1,2). Briefly, virus genome was reverse transcribed with multiple gene-specific primers targeting different regions of the viral genome. The synthesized cDNA was then subjected to multiple overlapping 2-kb PCRs for full-genome amplification. PCR amplicons obtained from the same specimen were pooled and sequenced using Novaseq or iSeq sequencing platform (Illumina). Specifically, for the sample from case-patient 1 with putative recombinant virus, we additionally performed NGS sequencing with the Illumina COVIDSeq kit (https://www.illumina.com/ products/ by-type/ivd-products/covidseq.html) for cross-validation. Generated sequencing reads were quality-trimmed by fastp (https://github.com/OpenGene/fastp) and mapped to a reference virus genome (Genbank accession: MN908947.3) by BWA-MEM2 v2.1 (3). Potential PCR duplicates were identified and removed by samtools markdup (https://www.htslib.org/doc/samtools-markdup.html. The genome consensus was generated by iVar with the PCR primer trimming protocol (minimum sequence depth of 5 for iSeq samples and minimum sequence depth of 10 for Novaseq samples, and minimum Q value of 30) (4). The deduced sequences can be available GISAID (Accession IDs: https://github.com/Leo-Poon-Lab/BA1_BA2_recombinant_HK/blob/main/GISAID_accessions.txt). We also cloned a ~2.2 kbp RT-PCR amplicon spanning the putative breakpoint region using Patient 2's sample. The 5' and 3' end of this clone was subjected to standard Sanger sequencing.

### *Identification of putative recombinants*

We scanned all the sequenced samples from imported cases in Hong Kong after November 15, 2021 for putative BA.1/BA.2 recombinants. The lineage defining mutations for BA.1 and BA.2 were curated from Cov-lineages (https://github.com/cov-lineages/pango-designation/issues/361) and CoVariants (https://covariants.org). For defining a sequence as a putative BA.1/BA.2 recombinant, it must have at least three BA.1- and BA.2- defining mutations, each with an allele frequency >90%. The statistics of sample's read depth, allele frequency and minor allele frequency were deduced from aligned reads in bam files using pysamstats (https://github.com/alimanfoo/pysamstats).

### *Identification of putative parental sequences of the recombinant*

The available public BA.1 and BA.2 sequences from GISAID and Genbank (accessed on 2022-03-07, totalling 1,222,642 and 767,399 entries, respectively) were downloaded and mapped to the reference sequence (Genbank accession: MN908947.3) using minimap2 (https://github.com/lh3/minimap2). The aligned sequences were used as the database for searching putative parental sequences. The leading/tailing partial sequences (positions 1 to 22005 and 21618 to 29903) of the recombinant genome were extracted by masking the remainder of the genome with "N"s. Masking was also performed for the aligned public sequences with letter "?" using figleaf (https://github.com/Koohoko/figleaf_fasta). The closest matches of the two partial recombinant sequences were separately identified from the above aligned public sequences using gofasta (https://github.com/virus-evolution/gofasta).

### *Phylogenetic analysis*

The consensus sequences deduced from NGS data were aligned to the reference genome (GenBank: MN908947.3) using MAFFT-add (https://mafft.cbrc.jp/alignment/server/add.html). The representative sequences from SARS-CoV-2 variants of concern Alpha, Beta, Gamma, Delta and Omicron BA.3 were also included. The 5' and 3' untranslated regions were masked before tree building. The Maximum likelihood phylogenies were estimated using IQ-TREE (v.2.1.3) (5), employing the best-fit nucleotide substitution models searched by the software with Wuhan-Hu-1 (GenBank: MN908947.3) as the outgroup. Branch supports are accessed by SH-aLRT and the ultrafast bootstrap, a node is considered supported if SH-aLRT >= 80% and UFboot >= 95% (http://www.iqtree.org/doc/Frequently-Asked-Questions).

### *Simplot analysis*

The putative BA.1/BA.2 recombinant virus sequence was analysed in Simplot v3.5.1 (6) for the recombination signals. Its similarity plot was plotted against a smaller group of representative variant of concerns (VOCs) including Alpha, Beta, Gamma, Delta, Omicron BA.1, Omicron BA.2, as well as the prototype Wuhan/WH01/2019 (Genbank accession: MN908947.3). Due to the relatively large proportion of strictly conserved sites, these sites were excluded from the alignment before subjecting to the similarity plot analysis. The BA.1/BA.2 recombinant, its putative parents from Omicron BA.1 and BA.2, and prototype Wuhan/WH01/2019 (as outgroup) were analysed for their informative sites of recombination.

The Genbank/GISAID accession numbers of viral sequences used in the Simplot analysis are as follow:

Alpha: OL807059, OU272361, OU052790, OU179605, OL315388, OU208527, OU174622, OU208088, MW933836, MZ280980, MZ296197, MZ077208, OU022681

Beta: OU202380, OU516338, OU136527, OK433425, OM765676, OL779105, OU114765, MW963525, OU233168

Gamma: MW913237, OL803729, MZ414874, MZ217960, MZ536412, OV921949, MZ211976, OM485550, MZ037589

Delta: OK208965, OK258803, MZ988451, OK101403, OK243904, MZ764878, OK160402, OK054978, MZ888548, MZ888540, MZ888535, MZ888534, OU338538, EPI_ISL_8880068

BA.1: EPI_ISL_10273412

BA.2: EPI_ISL_10462716

### *Code availability*

Detailed analysing scripts used in the study can be accessed in a GitHub repository (https://github.com/Leo-Poon-Lab/BA1_BA2_recombinant_HK/blob/main/GISAID_accessions.txt).

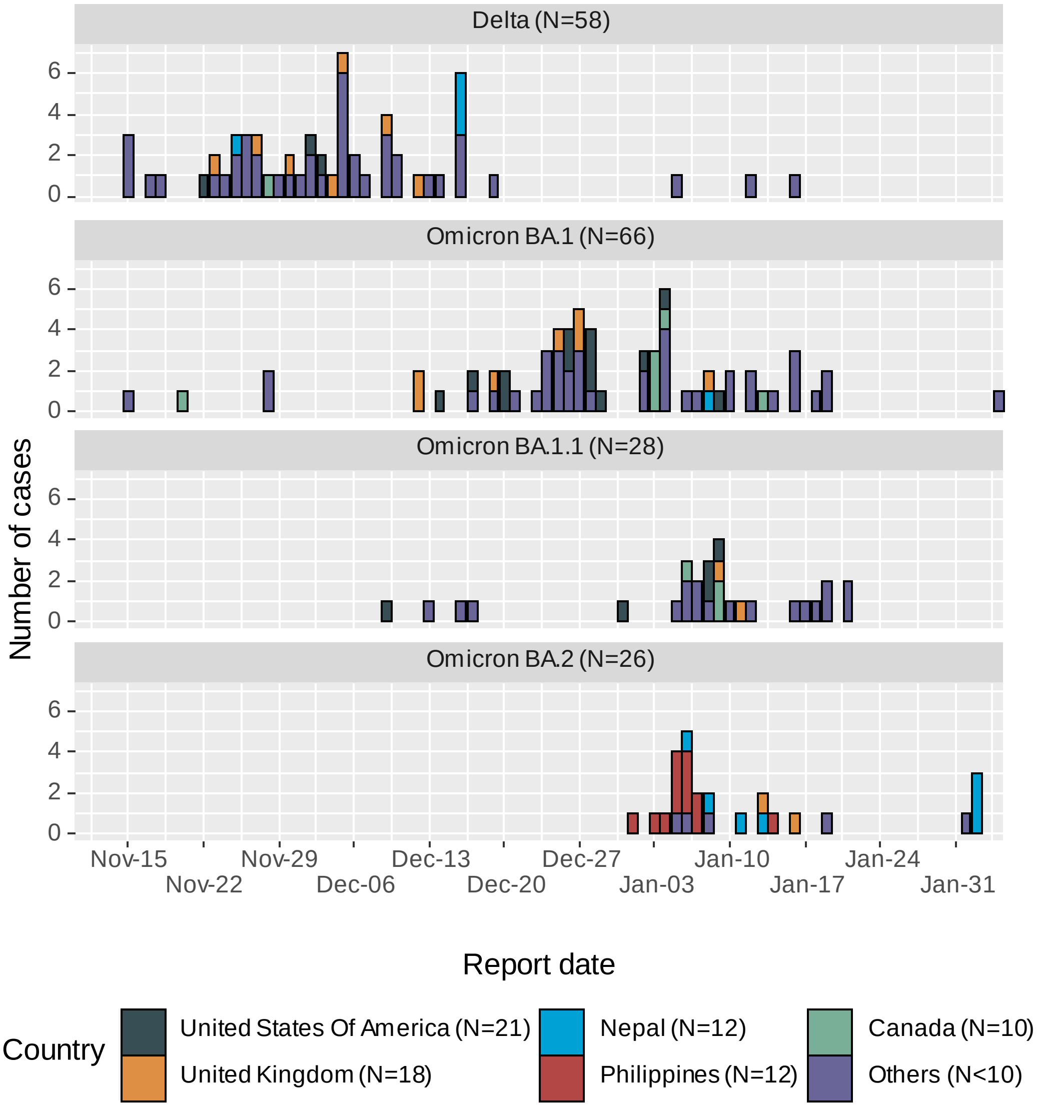


Appendix Figure 1. Importation of SARS-CoV-2 variants from incoming travellers. Dates of these patients first RT-PCR tested positive for SARS-CoV-2 and their countries of origin are as shown. All infections were confirmed by full-genome sequencing.

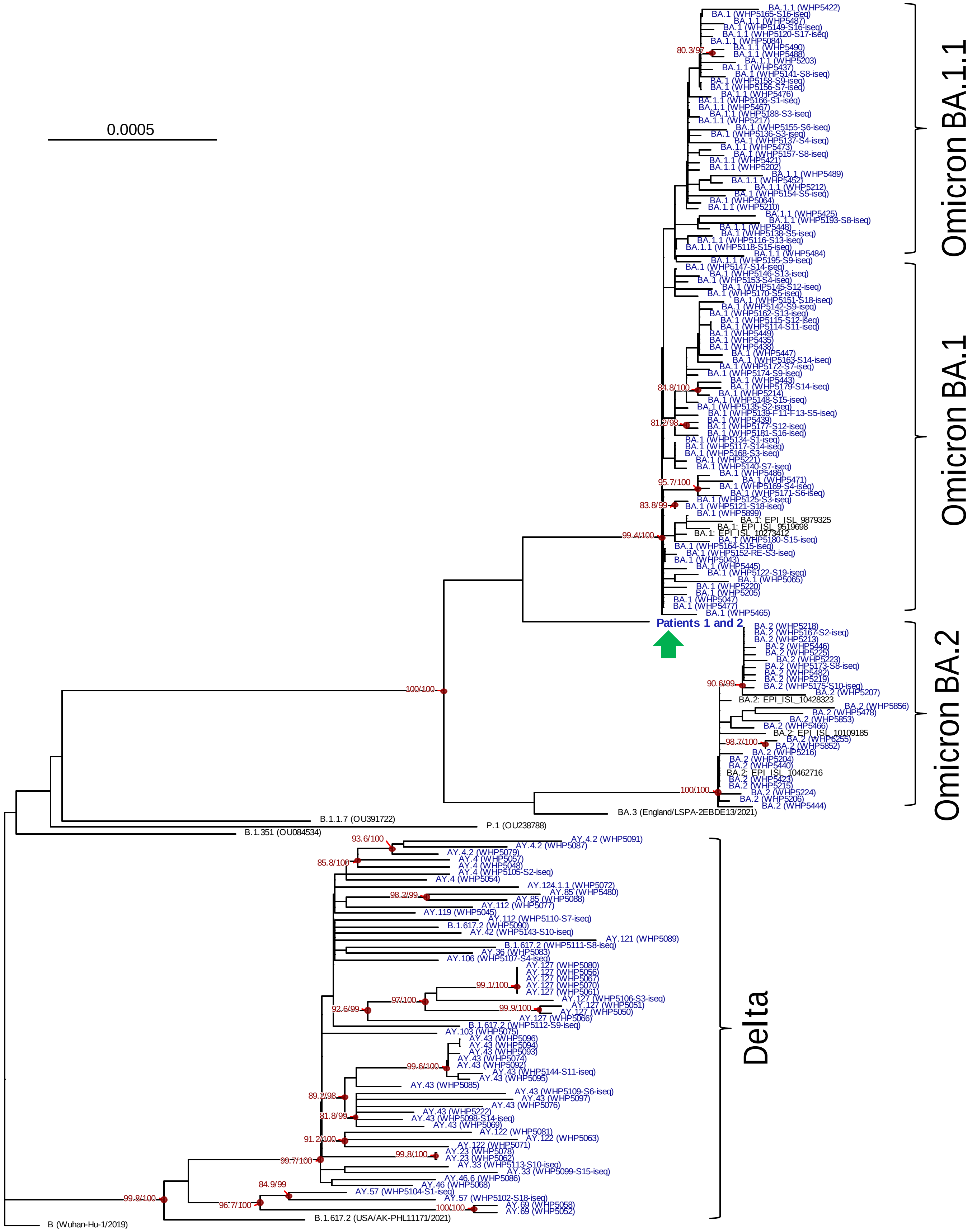


Appendix Figure 2. Phylogeny of SARS-CoV-2 variants identified in incoming travelers. The Maximum likelihood phylogenetic tree was generated using IQ-TREE employing the GTR+F+I nucleotide substitution model with Wuhan-Hu-1 (GenBank: MN908947.3) as the outgroup. Viral genomes generated from this study (blue) and references sequences used in the analysis are shown as indicated. The recombinant virus detected from Patients 1 and 2 is highlighted by an arrow. The red node points show strongly supported branches by SH-aLRT/ultrafast bootstrap. Scale bar indicates estimated genetic distance.

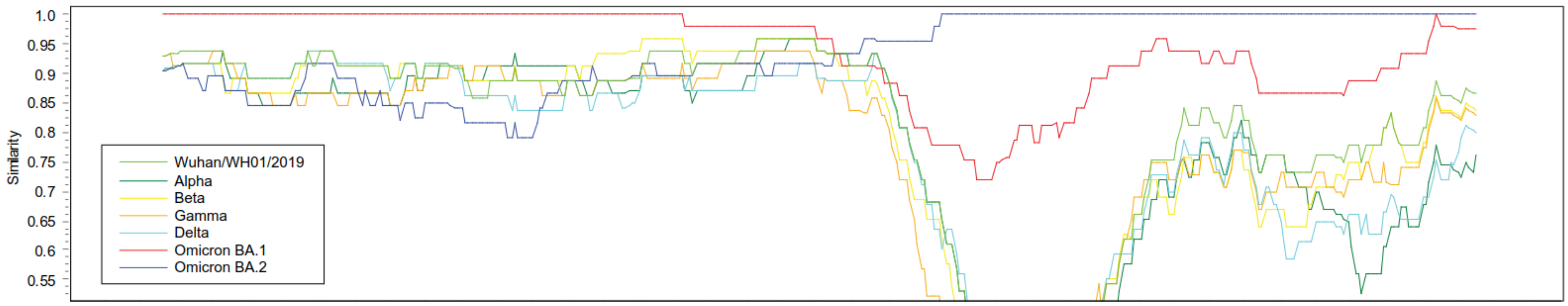


Appendix Figure 3. Simplot analysis of BA.1/BA.2 recombinant. Full viral genomes of different variant of concerns including Alpha, Beta, Gamma, Delta, Omicron BA.1, Omicron BA.2, as well as the prototype Wuhan/WH01/2019 were used in the analysis.

Appendix Table 1: Country of origin of imported COVID-19 cases

| Country of importation | N |
|---|---|
| United Kingdom | 23 |
| United States of America | 22 |
| Philippines | 13 |
| Nepal | 12 |
| Canada | 10 |
| Pakistan | 9 |
| Australia | 7 |
| Japan | 7 |
| Finland | 6 |
| France | 6 |
| India | 6 |
| Germany | 5 |
| Italy | 5 |
| Spain | 5 |
| Thailand | 5 |
| Ghana | 4 |
| Russia | 4 |
| Denmark | 3 |
| Ireland | 3 |
| Kenya | 3 |
| Switzerland | 3 |
| Vietnam | 3 |
| Brazil | 2 |
| Ethiopia | 2 |
| Kazakhstan | 2 |
| Korea | 2 |
| Nigeria | 2 |
| Poland | 2 |
| Singapore | 2 |
| South Africa | 2 |
| Sweden | 2 |
| Argentina | 1 |
| Bahamas | 1 |
| Belgium | 1 |
| Chile | 1 |
| Cyprus | 1 |
| Czech | 1 |
| Estonia | 1 |
| Lithuania | 1 |
| Morocco | 1 |
| Netherlands | 1 |
| Papua New Guinea | 1 |
| Qatar | 1 |
| Republic of Moldova | 1 |
| Saudi Arabia | 1 |
| Ukraine | 1 |
| United Republic of Tanzania | 1 |

**Appendix Table 2: GISAID sequences used in this study**

| Accession ID | Originating lab | Submitting lab | Authors |
|---|---|---|---|
| EPI_ISL_1996858, EPI_ISL_2146766 | Aegis Sciences Corporation | Centers for Disease Control and Prevention Division of Viral Diseases, Pathogen Discovery | Dakota Howard, Dhwani Batra, Peter W. Cook, Kara Moser, Adrian Paskey, Jason Caravas, Benjamin Rambo-Martin, Shatavia Morrison, Christopher Gulvick, Scott Sammons, Yvette Unoarumhi, Darlene Wagner, Matthew Schmerer, Cyndi Clark, Patrick Campbell, Rob Case, Vikramsinha Ghorpade, Holly Houdeshell, Ola Kvalvaag, Dillon Nall, Ethan Sanders, Alec Vest, Shaun Westlund, Matthew Hardison, Clinton R. Paden, Duncan MacCannell |
| EPI_ISL_3988521 | Colorado Department of Public Health and Environment | Colorado Department of Public Health and Environment | Laura Bankers, Molly C. Hetherington-Rauth, Diana Ir, Alexandria Rossheim, Michael Martin, Mandy Waters, Shannon R. Matzinger, Sarah Elizabeth Totten, Emily A. Travanty |
| EPI_ISL_3029243 | Department of Bacteria, Parasites and Fungi, Statens Serum Institut, Copenhagen, Denmark | Statens Serum Institut Bioinformatics and Microbial Genomics | Danish Covid-19 Genome Consortium |
| EPI_ISL_9519698 | ESPACEBIO | Department of Virology, Henri Mondor University Hospital, Assistance Publique Hôpitaux de Paris, Université Paris-Est Créteil, INSERM U955 | Christophe Rodriguez, Slim Fourati, Vanessa Demontant, Guillaume Gricourt, Melissa N'Debi, Alexandre Soulier, Elisabeth Trawinski, Jean-Michel Pawlotsky |
| EPI_ISL_1557096, EPI_ISL_3348121 | Fulgent Genetics | Centers for Disease Control and Prevention Division of Viral Diseases, Pathogen Discovery | Dakota Howard, Dhwani Batra, Peter W. Cook, Kara Moser, Adrian Paskey, Jason Caravas, Benjamin Rambo-Martin, Shatavia Morrison, Christopher Gulvick, Scott Sammons, Yvette Unoarumhi, Darlene Wagner, Matthew Schmerer, Harry Gao, Mickey Li, John Gao, Joseph Fierro, Benafsh Sapra, Becky Tsai, Yan Meng, Doreen Ng, James Xie, Clinton R. Paden, Duncan MacCannell |
| EPI_ISL_4359169, EPI_ISL_4454918 | Fulgent Genetics | Centers for Disease Control and Prevention Division of Viral Diseases, Pathogen Discovery | Dakota Howard,Dhwani Batra,Peter Cook,Jason Caravas,Benjamin Rambo-Martin,Scott Sammons,Yvette Unoarumhi,Matthew Schmerer,Kristine Lacek,Tymeckia Kendall,Victoria Caban Figueroa,Shatavia Morrison,Christopher Gulvick,Erisa Sula,Harry Gao,Mickey Li,John Gao,Joseph Fierro,Benafsh Sapra,Becky Tsai,Yan Meng,Doreen Ng,James Xie,Clinton Paden,Duncan MacCannell |
| EPI_ISL_4017432 | Fulgent Genetics | Centers for Disease Control and Prevention Division of Viral Diseases, Pathogen Discovery | Dakota Howard,Dhwani Batra,Peter Cook,Kara Moser,Adrian Paskey,Jason Caravas,Benjamin Rambo-Martin,Shatavia Morrison,Christopher Gulvick,Scott Sammons,Yvette Unoarumhi,Darlene Wagner,Matthew Schmerer,Harry Gao,Mickey Li,John Gao,Joseph Fierro,Benafsh Sapra,Becky Tsai,Yan Meng,Doreen Ng,James Xie,Clinton Paden,Duncan MacCannell |
| EPI_ISL_2614026 | Gravity Diagnostics, LLC | Gravity Diagnostics, LLC | Gravity Diagnostics |

| | | | |
|---|---|---|---|
| EPI_ISL_9879325 | Hopital | National Reference Center for Viruses of Respiratory Infections, Institut Pasteur, Paris | Marion Barbet, Sylvie Behillil, Méline Bizard, Angela Brisebarre, Camille Capel, Vincent Enouf, Louise Lefrançois, Frédéric Lemoine, Christophe Malabat, Corinne Maufrais, Slim El Khiari, Julien Fumey, Etienne Simon-Lorière, Maud Vanpeene, Sylvie Van der Werf, Benedicte LUREAU |
| EPI_ISL_2282971 | Infinity Biologix | Centers for Disease Control and Prevention Division of Viral Diseases, Pathogen Discovery | Dakota Howard, Dhwani Batra, Peter W. Cook, Kara Moser, Adrian Paskey, Jason Caravas, Benjamin Rambo-Martin, Shatavia Morrison, Christopher Gulvick, Scott Sammons, Yvette Unoarumhi, Darlene Wagner, Matthew Schmerer, Christian Bixby, Yihe Wang, Jonathan Schultz, Chirayu Goswami, Russ Hager, Robin Grimwood, Clinton R. Paden, Duncan MacCannell |
| EPI_ISL_1682200 | Laboratory Corporation of America | Centers for Disease Control and Prevention Division of Viral Diseases, Pathogen Discovery | Dakota Howard, Dhwani Batra, Peter W. Cook, Kara Moser, Adrian Paskey, Jason Caravas, Benjamin Rambo-Martin, Shatavia Morrison, Christopher Gulvick, Scott Sammons, Yvette Unoarumhi, Darlene Wagner, Matthew Schmerer, Minoo Agarwal, Eyad Almasri, Debbie Boles, Ayla Burns, Nuthawin Charoensri, Oren Cohen, Susan Countryman, Mary Ann Cristobal, Bobbi Croy, Suzanne Dale, Hrushikesh Deshmukh, Amanda Douglas, Vincent Drouillon, Marcia Eisenberg, Howard Engler, Rama Ghatti, Prashant Gupta, Susan Hicks, Jake Humphrey, Lax Iyer, Manoj Jain, Mohan Kolli, Brian Krueger, Tim Kuphal, Stanley Letovsky, Michael Levandoski, Craig Lukasik, Jonathan Meltzer, Brian Norvell, Mindy Nye, Scott Parker, Christos Petropoulos, John Pruitt, Steven Ragan, Scott Ryan, Mike Sapeta, Jana Schroth, Suresh Babu Selvaraju, Goran Stevovic, Amanda Suchanek, Andrea Throop, Lyndon Tilson, Thomas Urban, Joe Voshell, Kimberly Wagner, Jonathan Williams, Mary Williamson, Qian Zeng, Tricia Zwiefelhofer, Clinton R. Paden, Duncan MacCannell |
| EPI_ISL_10273412 | Laboratory Corporation of America | Centers for Disease Control and Prevention Division of Viral Diseases, Pathogen Discovery | Dakota Howard,Dhwani Batra,Peter Cook,Jason Caravas,Benjamin Rambo-Martin,Scott Sammons,Yvette Unoarumhi,Matthew Schmerer,Kristine Lacek,Tymeckia Kendall,Victoria Caban Figueroa,Shatavia Morrison,Christopher Gulvick,Minoo Agarwal,Eyad Almasri,Debbie Boles,Ayla Burns,Nuthawin Charoensri,Oren Cohen,Susan Countryman,Mary Cristobal,Bobbi Croy,Suzanne Dale,Hrushikesh Deshmukh,Amanda Douglas,Vincent Drouillon,Marcia Eisenberg,Howard Engler,Rama Ghatti,Prashant Gupta,Susan Hicks,Jake Humphrey,Lax Iyer,Lisa Pfefferle,Manoj Jain,Matthew Robinson,Mohan Kolli,Brian Krueger,Tim Kuphal,Stanley Letovsky,Michael Levandoski,Craig Lukasik,Jonathan Meltzer,Brian Norvell,Mindy Nye,Scott Parker,Christos Petropoulos,John Pruitt,Steven Ragan,Scott Ryan,Mike Sapeta,Jana Schroth,Suresh Selvaraju,Goran Stevovic,Amanda Suchanek,Andrea Throop,Lyndon Tilson,Thomas Urban,Joe Voshell,Kimberly Wagner,Jonathan Williams,Mary Williamson,Qian Zeng,Tricia Zwiefelhofer,Clinton Paden,Duncan MacCannell |
| EPI_ISL_3745736, EPI_ISL_4153131 | Laboratory Corporation of America | Centers for Disease Control and Prevention Division of Viral Diseases, Pathogen Discovery | Dakota Howard,Dhwani Batra,Peter Cook,Kara Moser,Adrian Paskey,Jason Caravas,Benjamin Rambo-Martin,Shatavia Morrison,Christopher Gulvick,Scott Sammons,Yvette Unoarumhi,Darlene Wagner,Matthew Schmerer,Minoo Agarwal,Eyad Almasri,Debbie Boles,Ayla Burns,Nuthawin Charoensri,Oren Cohen,Susan Countryman,Mary Cristobal,Bobbi Croy,Suzanne Dale,Hrushikesh Deshmukh,Amanda |

| | | | |
|---|---|---|---|
| | | | Douglas,Vincent Drouillon,Marcia Eisenberg,Howard Engler,Rama Ghatti,Prashant Gupta,Susan Hicks,Jake Humphrey,Lax Iyer,Lisa Pfefferle,Manoj Jain,Matthew Robinson,Mohan Kolli,Brian Krueger,Tim Kuphal,Stanley Letovsky,Michael Levandoski,Craig Lukasik,Jonathan Meltzer,Brian Norvell,Mindy Nye,Scott Parker,Christos Petropoulos,John Pruitt,Steven Ragan,Scott Ryan,Mike Sapeta,Jana Schroth,Suresh Selvaraju,Goran Stevovic,Amanda Suchanek,Andrea Throop,Lyndon Tilson,Thomas Urban,Joe Voshell,Kimberly Wagner,Jonathan Williams,Mary Williamson,Qian Zeng,Tricia Zwiefelhofer,Clinton Paden,Duncan MacCannell |
| EPI_ISL_10462716, EPI_ISL_1454544, EPI_ISL_2021385 | Lighthouse Lab in Alderley Park | Wellcome Sanger Institute for the COVID-19 Genomics UK (COG-UK) Consortium | Jacquelyn Wynn, Mairead Hyland, The Lighthouse Lab in Alderley Park and Alex Alderton, Roberto Amato, Jeffrey Barrett, Sonia Goncalves, Ewan Harrison, David K. Jackson, Ian Johnston, Dominic Kwiatkowski, Cordelia Langford, John Sillitoe on behalf of the Wellcome Sanger Institute COVID-19 Surveillance Team |
| EPI_ISL_1518044 | Lighthouse Lab in Cambridge | Wellcome Sanger Institute for the COVID-19 Genomics UK (COG-UK) Consortium | Rob Howes, The Lighthouse Lab in Cambridge and Alex Alderton, Roberto Amato, Jeffrey Barrett, Sonia Goncalves, Ewan Harrison, David K. Jackson, Ian Johnston, Dominic Kwiatkowski, Cordelia Langford, John Sillitoe on behalf of the Wellcome Sanger Institute COVID-19 Surveillance Team |
| EPI_ISL_1115472, EPI_ISL_1256650, EPI_ISL_1410322, EPI_ISL_1453906, EPI_ISL_2608121 | Lighthouse Lab in Glasgow | Wellcome Sanger Institute for the COVID-19 Genomics UK (COG-UK) Consortium | Harper VanSteenhouse, Yumi Kasai, David Gray, Carol Clugston, Anna Dominiczak and Alex Alderton, Roberto Amato, Jeffrey Barrett, Sonia Goncalves, Ewan Harrison, David K. Jackson, Ian Johnston, Dominic Kwiatkowski, Cordelia Langford, John Sillitoe on behalf of the Wellcome Sanger Institute COVID-19 Surveillance Team |
| EPI_ISL_10109185, EPI_ISL_10428323 | Lighthouse Lab in Milton Keynes | Wellcome Sanger Institute for the COVID-19 Genomics UK (COG-UK) Consortium | The Lighthouse Lab in Milton Keynes and Alex Alderton, Roberto Amato, Jeffrey Barrett, Sonia Goncalves, Ewan Harrison, David K. Jackson, Ian Johnston, Dominic Kwiatkowski, Cordelia Langford, John Sillitoe on behalf of the Wellcome Sanger Institute COVID-19 Surveillance Team |
| EPI_ISL_2959430, EPI_ISL_3129581 | Michigan Department of Health and Human Services, Bureau of Laboratories | Michigan Department of Health and Human Services, Bureau of Laboratories | Blankenship HM, Riner D, Soehnlen MK |
| EPI_ISL_1016848 | North Shore Hospital | Institute of Environmental Science and Research (ESR) | Xiaoyun Ren, Matt Storey, Nikki Freed, Muhammad Faisal, Jing Wang, Hermes Perez, Anja Werno, Antje van der Linden, Arlo Upton, Chris Mansell, David Hammer, Dragana Drinkovic, Gary McAuliffe, Hana Sofia Andersson, James Ussher, Jill Sherwood, Josh Freeman, Julia Howard, Juliet Elvy, Mary DeAlmeida, Matt Blakiston, Matthew Rogers, Max Bloomfield, Michael Addidle, Michelle Balm, Sally Roberts, Sarah Jefferies, Sharmini Muttaiyah, Susan Morpeth, Susan Taylor, Timothy Blackmore, Vani Sathyendran, Veronica Playle, Virginia Hope, Erasmus Smit, Lauren Jelly, Olin Silander, Joep de Ligt |
| EPI_ISL_2876740 | Quest Diagnostics Incorporated | Centers for Disease Control and Prevention Division of Viral | Dakota Howard, Dhwani Batra, Peter W. Cook, Kara Moser, Adrian Paskey, Jason Caravas, Benjamin Rambo-Martin, Shatavia Morrison, Christopher Gulvick, Scott Sammons, Yvette Unoarumhi, Darlene Wagner, Matthew Schmerer, S. H. |

| | | | |
|---|---|---|---|
| | | Diseases, Pathogen Discovery | Rosenthal, A. Gerasimova, R. M. Kagan, B. Anderson, M. Hua, Y. Liu, L.E. Bernstein, K.E. Livingston, A. Perez, I. A. Shlyakhter, R. V. Rolando, R. Owen, P. Tanpaiboon, F. Lacbawan, Clinton R. Paden, Duncan MacCannell |
| EPI_ISL_2133917 | Rhode Island Department of Health | Infectious Disease Program, Broad Institute of Harvard and MIT | Siddle,K.J., Azevedo,K., Miller,A., Adams,G., Pearlman,L., Gladden-Young,A., Lagerborg,K., Rudy,M., DeRuff,K., Carter,A., Normandin,E., Bauer,M., Reilly,S., Tomkins-Tinch,C., Loreth,C., Chaluvadi,S., Lemieux,J.E., Birren,B.W., Sabeti,P.C., Huard,R., King,E., Park,D.J., and MacInnis,B.L. |
| EPI_ISL_8880068 | Temporary Specimen Collection Centre at the AsiaWorld-Expo | Hong Kong Department of Health | Alan K.L. Tsang, Peter C.W. Yip, Patricia K. L. Leung, Ken H.L. Ng, Edman T.K. Lam, Rickjason C.W. Chan |
| EPI_ISL_3695744 | TPMG Regional Laboratory | California Department of Public Health | Emily Smith on behalf of CDPH-COVIDNet and UCI Genome Sciences Center/GHTF |
| EPI_ISL_3409895 | UMass Memorial Medical Center | Center for Microbiome Research | Richard T. Ellison III, Karl J. Simon, Doyle V. Ward |
| EPI_ISL_3506433 | Victoria Hospital wc VHW | NHLS/UCT | Arash Iranzadeh, Deelan Doolabh, Lynn Tyers, Bruna Galvao, Innocent Mudau, Marvin Hsiao, Gert Marais, Diana Hardie, Stephen Korsman, Rageema Joseph, Carolyn Williamson |
| EPI_ISL_2151915 | Viollier AG | Department of Biosystems Science and Engineering, ETH Zürich | Chaoran Chen, Sarah Nadeau, Catharine Aquino, Ivan Topolsky, Philipp Jablonski, Lara Fuhrmann, David Dreifuss, Katharina Jahn, Daniel Ehrsam, Isabel Stürmer, Andreia Cabral de Gouvea, Maria Domenica Moccia, Simon Grüter, Timothy Sykes, Lennart Opitz, Griffin White, Laura Neff, Doris Popovic, Andrea Patrignani, Jay Tracy, Ralph Schlapbach, Christiane Beckmann, Maurice Redondo, Olivier Kobel, Christoph Noppen, Sophie Seidel, Noemie Santamaria de Souza, Niko Beerenwinkel, Tanja Stadler |

We gratefully acknowledge the following Authors from the Originating laboratories responsible for obtaining the specimens and the Submitting laboratories where genetic sequence data were generated and shared via the GISAID Initiative, on which this research is based.
